# Untargeted metabolomics profiling in pediatric patients and adult populations indicates a connection between lipid imbalance and epilepsy

**DOI:** 10.1101/2023.03.29.23287640

**Authors:** Kaisa Teele Oja, Mihkel Ilisson, Karit Reinson, Kai Muru, Tiia Reimand, Hedi Peterson, Dmytro Fishman, Tõnu Esko, Toomas Haller, Jaanika Kronberg, Monica H. Wojcik, Adam Kennedy, Gregory Michelotti, Anne O’Donnell-Luria, Eve Õiglane-Šlik, Sander Pajusalu, Katrin Õunap

## Abstract

**Introduction:** Epilepsy is a common central nervous system disorder characterized by abnormal brain electrical activity. We aimed to compare the metabolic profiles of plasma from patients with epilepsy across different etiologies, seizure frequency, seizure type, and patient age to try to identify common disrupted pathways.

**Material and methods:** We used data from three separate cohorts. The first cohort (PED-C) consisted of 31 pediatric patients with suspicion of a genetic disorder with unclear etiology; the second cohort (AD-C) consisted of 250 adults from the Estonian Biobank (EstBB), and the third cohort consisted of 583 adults ≥ 69 years of age from the EstBB (ELD-C). We compared untargeted metabolomics and lipidomics data between individuals with and without epilepsy in each cohort.

**Results:** In the PED-C, significant alterations (p-value <0.05) were detected in sixteen different glycerophosphatidylcholines (GPC), dimethylglycine and eicosanedioate (C20-DC). In the AD-C, nine significantly altered metabolites were found, mainly triacylglycerides (TAG), which are also precursors in the GPC synthesis pathway. In the ELD-C, significant changes in twenty metabolites including multiple TAGs were observed in the metabolic profile of participants with previously diagnosed epilepsy. Pathway analysis revealed that among the metabolites that differ significantly between epilepsy-positive and epilepsy-negative patients in the PED-C, the lipid superpathway (p = 3.2*10-4) and phosphatidylcholine (p = 9.3*10-8) and lysophospholipid (p = 5.9*10-3) subpathways are statistically overrepresented. Analogously, in the AD-C, the triacylglyceride subclass turned out to be statistically overrepresented (p = 8.5*10-5) with the lipid superpathway (p = 1.4*10-2). The presented p-values are FDR-corrected.

**Conclusion:** Our results suggest that cell membrane fluidity may have a significant role in the mechanism of epilepsy, and changes in lipid balance may indicate epilepsy. However, further studies are needed to evaluate whether untargeted metabolomics analysis could prove helpful in diagnosing epilepsy earlier.

## Introduction

Epilepsy is a common central nervous system disorder characterized by abnormal brain electrical activity and defined as having at least two unprovoked seizures or one unprovoked seizure with a high probability of further seizures [^1^]. An estimated 50 million individuals worldwide have been diagnosed with epilepsy [^2^]. It can occur among children and adults, with the highest incidence generally among the youngest and oldest age groups, although the etiology differs [^3–6^]. In a recent epidemiological study of childhood epilepsy in Estonia the overall incidence was 86.3 cases per 100 000 person-years, however the highest incidence was among children between 5-9 years of age [^7^]. A demonstrable cause is more likely among patients with early-onset epilepsy, with genetic and structural causes being the most common [^5,7^]. In patients over 60 years of age, the leading cause of epilepsy is stroke [^6^]. In Estonia, the reported prevalence rate (PR) of childhood epilepsy is 3.6 per 1000 [^8^], and the PR of epilepsy in the adult population is 5.3 per 1000 population [^9^], which is comparable to prevalence rates reported in other developed countries [^3^]. About a third of patients have drug-resistant epilepsy and poor seizure control [^10^].

The classification system of epilepsy is complex and places great importance on ascertaining the etiology [^11^]. Different metabolic disorders are associated with epilepsy and constitute a separate etiological group [^9^]. However, Wang et al. showed that common metabolic features could be found in at least three types of seizures [^12^]. Altered levels of glutamate, lactate, and citrate have been associated with epilepsy in addition to changes in multiple metabolic pathways like alanine, aspartate and glutamate metabolism, glycine, serine and threonine metabolism, glycerophospholipid metabolism, arginine and proline metabolism [^13^]. Recent studies have therefore focused on metabolomics to understand the pathophysiology of epilepsy and find new targets for antiepileptic drugs (AEDs) [^12,13^].

We tested the hypothesis that there are similarities in the metabolic profiles of patients with epilepsy despite different etiology, seizure frequency, seizure type, and patient age via untargeted metabolomics and lipidomics analysis of individuals with and without epilepsy in three cohorts.

## Materials & Methods

### Ethics

The study was conducted in accordance with the Declaration of Helsinki, and approval was granted by the Research Ethics Committee of the University of Tartu (Certificates No 259/T-2, 263/M-16, 283/M-10, 287/M-15 and 374/M-8) and the Estonian Committee on Bioethics and Human Research (Certificate No 1.1-12/3749). Informed consent was obtained from all individuals or their legal guardians before enrollment in the study. Participants aged 7-17 were also given an age-appropriate informed consent form.

### Data availability

All analyzed data consists of patient’s personal data and is stored according to regulations of the institutions. Pseudonymized data is available on request.

### Subjects

We used data from three cohorts that varied regarding general characteristics (**Table 1**) due to different inclusion criteria. The first cohort (PED-C) consisted of 31 pediatric patients with suspicion of a genetic disorder with unclear etiology enrolled from the Department of Clinical Genetics at Tartu University Hospital between 2016-2018; the second cohort (AD-C) consisted of 250 adults from the Estonian Biobank (EstBB) [^14^], and the third cohort (ELD-C) consisted of 583 elderly participants from the EstBB [^15^]. The EstBB is a volunteer-based biobank representing about 5% of the Estonian adult population [^16^]. All participants from the EstBB had been enrolled before 2011, and the primary criterion for inclusion in this project was the availability of suitable metabolomics data.

**Table 1.** General characteristics of all three cohorts. The mean age of onset was calculated for epilepsy-positive subgroups in each cohort.

| Characteristic | PED-C<br>(n=31) | AD-C<br>(n=250) | ELD-C<br>(n=583) |
| --- | --- | --- | --- |
| Mean (range) age in years | 9 (2-17) | 39 (20-64) | 73 (69-79) |
| Gender |  |  |  |
| Male (%) | 20 (64.5) | 128 (51.2) | 176 (30.2) |
| Epilepsy diagnosis (%) | 11 (35.5) | 11 (4.4) | 26 (4.5) |
| Mean (range) age of<br>epilepsy onset in years | 4.4 (0.17-15) | 28.8 (5-64) | 79.4 (66-90) |

For most patients in the PED-C, no molecular diagnosis could be confirmed via testing through routine clinical practice (diagnosed with ORPHA:616874 code – rare disorder without a determined diagnosis after full investigation). Two exceptions were made – one patient had an O-linked N-acetylglucosamine transferase (OGT) defect with a confirmed molecular diagnosis but was included as a control, and another patient had been diagnosed with xanthinuria but this diagnosis did not explain the entire phenotype. All the patients in the PED-C were included in our research project in collaboration with the Center for Mendelian Genomics (CMG) at the Broad Institute of MIT and Harvard.

The AD-C consisted of participants from a larger cohort selected to represent body-mass-index (BMI) extremes and their randomly matched controls [^**14**^]. Individuals with pregnancy, anorexia, or a known wasting illness were excluded [^**14**^]. The original cohort consisted of 300 individuals, with an equal number of the lean, obese, and general population from the EstBB [^**14**,**17**^]. Our subset included 100 population controls, 73 lean and 77 obese individuals. We used only part of the original cohort for our analysis due to technical issues during data transfer.

The ELD-C included participants aged between 69 and 79 years at the time of sampling, and individuals with a preexisting history of hypertensive heart disease, diabetes, coronary artery disease, cancer, chronic obstructive pulmonary disease, stroke, or Alzheimer’s disease were excluded [^15^]. The two EstBB cohorts do not overlap [^14^].

We requested access to the metabolomics and lipidomics data of the AD-C and ELD-C, which had already been analyzed in separate projects [^**14**,**15**,**17**^]. The Estonian Committee on Bioethics and Human Research approved our request. The activities of the EstBB are regulated by the Human Genes Research Act [^**18**^], which was adopted in 2000 specifically for the operations of the EstBB. Individual level data analysis in the EstBB was carried out under ethical approval [ethics approval no 1.1-12/3749 from the Estonian Committee on Bioethics and Human Research (Estonian Ministry of Social Affairs), using data according to release application “R04”] from the Estonian Biobank. Informed consent had been obtained from all participants before joining the EstBB to use their biological samples for future studies.

### Clinical data collection

In the PED-C, electronic medical records were analyzed, mainly from the Genetics and Personalized Medicine Clinic and the Children’s Clinic of Tartu University Hospital. Information on the patients’ intellect, behavior, development, growth, facial features, epilepsy, brain structure, and movement was collected in addition to general characteristics such as age, gender, and perinatal history. In the AD-C and the ELD-C, information on gender, age, body mass index (BMI), diagnosis of dyslipidemia and epilepsy, age at epilepsy diagnosis, treatment with antiepileptic drugs (Anatomical Therapeutic Chemical (ATC) code N03A) at the time of sampling was requested from the EstBB. Dyslipidemia was defined as the diagnosis of E78 (disorders of lipoprotein metabolism and other lipidemias) according to the International Classification of Diseases (ICD-10), and epilepsy as G40 (epilepsy) or G41 (status epilepticus) with all their subgroups [^19^].

### Metabolomics analyses

In the PED-C, 4 ml of EDTA plasma was collected from all patients for metabolomics and lipidomics analysis. Fasting overnight before sampling was recommended but not a strict requirement. All patients had fasted for at least three hours, and eleven had fasted overnight. The collected plasma samples were frozen (≤ −20°C) and shipped out to Metabolon Inc in the USA [^20^], where untargeted metabolomics and lipidomics analysis was performed using the Meta IMD test. This Laboratory-Developed Test (LDT), performed in a CLIA-certified laboratory, detects over 800 small molecules (ranging from 50 to 1 500 Daltons in molecular weight). The test uses four different methods of high-performance Ultra Performance Liquid Chromatography (UPLC) instruments paired with Mass Spectrometry (UPLC/ MS). Detected molecules were identified by comparison to a chemical reference library, with unique biochemical entries characterized by accurate molecular mass, including information on any adductation, in source fragmentation, and/or polymerization (typically dimers and trimers) and retention time/index on the chromatography columns. The results were reported as z-scores calculated using a reference cohort of 866 pediatric samples. Individual concentrations were not reported.

In the AD-C, metabolite profiling had also been done using four liquid chromatography-mass spectrometry (LC-MS) methods [^14^]. We received data about 327 known metabolites and 17,774 unknown signals.

The ELD-C non-fasting plasma samples were collected and stored in liquid nitrogen before shipment to the Broad Institute (Cambridge, MA, USA) for metabolomic profiling using LC-MS [^15^]. The metabolites were extracted with four different methods described in detail by Esko et al., 2017 [^21^]. We received data about 585 known metabolites and >19,000 unknown signals. The methods used for the AD-C and ELD-C have been described in detail by Hsu et al. in 2019 [^14^].

### Statistical testing

The computational and programming tasks were performed on a personal computer with Python 3.7. Pandas, Numpy, Sklearn, and Scipy libraries were used for data handling and statistical testing.

Statistical tests were done separately in each cohort.

Univariate statistical tests were performed to investigate whether a particular metabolite could be associated with epilepsy. Kruskal-Wallis tests were run, and after confirming the normal metabolite distributions for both epilepsy positive and negative classes by Shapiro-Wilk test (p > 0.05), Welch t-tests were also run. Extracted p-values for each test were collected into two individual files and corrected with FDR using the Benjamini-Hochberg approach. For interpretation of results, significance threshold α = 0.05 was used.

Pathway overrepresentation analyses were evaluated by hypergeometric testing using a significance threshold α = 0.05. P-values were corrected with FDR using the Benjamini-Hochberg approach.

## Results

### Clinical features

In the PED-C, all patients were white European, the male to female ratio was 20:11, and eleven (35.5%) patients had been diagnosed with epilepsy. In seven cases, the diagnosis was G40.2 Localization-related (focal, partial) symptomatic epilepsy and epileptic syndromes with complex partial seizures; others were diagnosed with G40.3 (Generalized idiopathic epilepsy and epileptic syndromes), G40.4 (Other generalized epilepsy and epileptic syndromes) and/or G41.9 (Status epilepticus, unspecified). For epilepsy-positive patients, we also collected data about the age of onset, age at diagnosis, and treatment at the time of sampling (**Table S1**). All cases were diagnosed before metabolomics sampling, and two patients did not receive any medication at the time. Two patients used one, two patients used two, and five patients had a combination of three or more drugs. The most common treatment was with lamotrigine (four patients); other medications, including levetiracetam, valproic acid, and vigabatrin, were all given to two patients. None of the patients were on a ketogenic diet. The most common comorbidities were developmental delay (24/31, 77.4%), intellectual disability (21/31, 67.7%), and dysmorphic features (20/31, 64.5%). There were no statistically significant differences in age, gender, or frequency of comorbidities between epilepsy-positive and negative subgroups in the PED-C. However, intellectual disability was present in almost all epilepsy patients and only in about half of the patients without epilepsy (**Table 2**). The only exception in the epilepsy subgroup is an individual with borderline intellectual disability; in 2019, exome sequencing reanalysis revealed a novel *de novo* frameshift variant in *CSNK2A1* that was considered to be likely pathogenic by ACMG classification [^22^]. He has been seizure free for the past three years. The patient with a known OGT defect belonged to the epilepsy-negative subgroup, while the patient with xanthinuria and an unknown disorder had epilepsy.

**Table 2.** Characteristics of epilepsy positive and negative subgroups in the pediatric cohort. Fisher’s exact test was used for categorical variables, and the Welch t-test was used for continuous variables.

| Clinical features in the PED-C | Patients with epilepsy (n=11) | Patients without epilepsy (n=20) | p-value |
| --- | --- | --- | --- |
| Mean age (range) | 11 (3-17) | 8.45 (2-14) | 0.15 |
| Female (%) | 5 (45.5) | 6 (70) | 0.45 |
| Intellectual disability (%) | 10 (90.9) | 11 (55) | 0.06 |
| Neuromuscular disorders (%) | 8 (72.7) | 9 (45) | 0.26 |
| Developmental delay (%) | 10 (90.9) | 14 (70) | 0.37 |
| Dysmorphic features (%) | 6 (54.5) | 14 (70) | 0.45 |
| Brain malformations (%) | 7 (63.6) | 10 (50) | 0.71 |
| Autism spectrum disorder (%) | 5 (45.5) | 9 (45) | 1 |
| Abnormal growth (%) | 5 (45.5) | 9 (45) | 1 |
| Microcephaly (%) | 4 (36.3) | 6 (30) | 1 |

In the EstBB cohorts, epilepsy was present in 2.8% of the AD-C and 0.9% of the ELD-C at the time of sampling. The prevalence in the ELD-C is similar to other reports in this age group in developed countries [^23^]. However, the age-adjusted (to the 1970 US population) prevalence rate of epilepsy in the Estonian adult population is lower than the prevalence we observed in the AD-C [^9^]. At the time of data retrieval, 25 additional cases had been diagnosed, and the final prevalence was 4.4% in the AD-C and 4.5% in the ELD-C. At the time of metabolome sampling, four participants with epilepsy were receiving treatment with antiepileptic drugs (respectively carbamazepine, primidone, topiramate, and clonazepam). Dyslipidemia had been diagnosed in 288 cases out of 833 (34.57%) in the EstBB cohorts – 65 cases (26%) in the AD-C and 223 (38.3%) in the ELD-C. Among participants with epilepsy, dyslipidemia diagnosis was present in 13 cases (35.14%) at the time of data retrieval. It was distributed evenly between the two cohorts – four cases (36.4%) in the AD-C and nine cases (34.6%) in the ELD-C. The mean age, gender distribution, and BMI did not differ statistically significantly between epilepsy positive and negative subgroups in either EstBB cohort (**Table 3**).

**Table 3.** Characteristics of the AD-C and ELD-C subgroups based on the presence of epilepsy. The Welch t-test was used for age; the chi-square test for gender; the Wilcoxon rank sum test with continuity correction for BMI; and Fisher’s exact test for dyslipidemia diagnosis. Statistical significance was defined as p-value < 0.05.

| Clinical features | AD-C |  |  | ELD-C |  |  |
| --- | --- | --- | --- | --- | --- | --- |
|  | Epilepsy (n=11) | No epilepsy (n=239) | p-value | Epilepsy (n=26) | No epilepsy (n=557) | p-value |
| Mean age (range) | 35 (20-56) | 39 (20-64) | 0.36 | 73 (70-78) | 73 (69-79) | 0.61 |
| Female (%) | 3 (27.3) | 119 (49.8) | 0.25 | 14 (53.8) | 393 (70.5) | 0.11 |
| Median (IQR) BMI | 24.3 (7.5) | 26.4 (17.7) | 0.36 | 27.7 (7.5) | 27.0 (5.4) | 0.97 |
| Dyslipidemia diagnosis (%) | 4 (36.4) | 61 (25.5) | 0.48 | 9 (34.6) | 214 (38.4) | 0.83 |
| Epilepsy onset before sampling (%) | 7 (63.6) | NA | NA | 5 (19.2) | NA | NA |

### Metabolomics results

In the PED-C, the levels of eighteen metabolites out of 709 were significantly different (FDR-corrected p-value <0.05) between patients with and without epilepsy. These included sixteen different glycerophosphatidylcholines (GPC), dimethylglycine and eicosanedioate (C20-DC) (**Table S2**). In the AD-C, nine significantly altered metabolites were found, mainly triacylglycerides (TAG), which are also precursors in the GPC synthesis pathway, and one phosphatidylethanolamine (**Table S2**). The only significantly altered metabolite in the ELD-C was hydrochlorothiazide, with an average Z-score of −0.2145 (corrected p-value <0.001). In the EstBB cohorts, all the altered metabolites were lower than average in the epilepsy subgroup, but in the PED-C, some GPCs were increased instead (**Table 4, Table S2**).

**Table 4.**
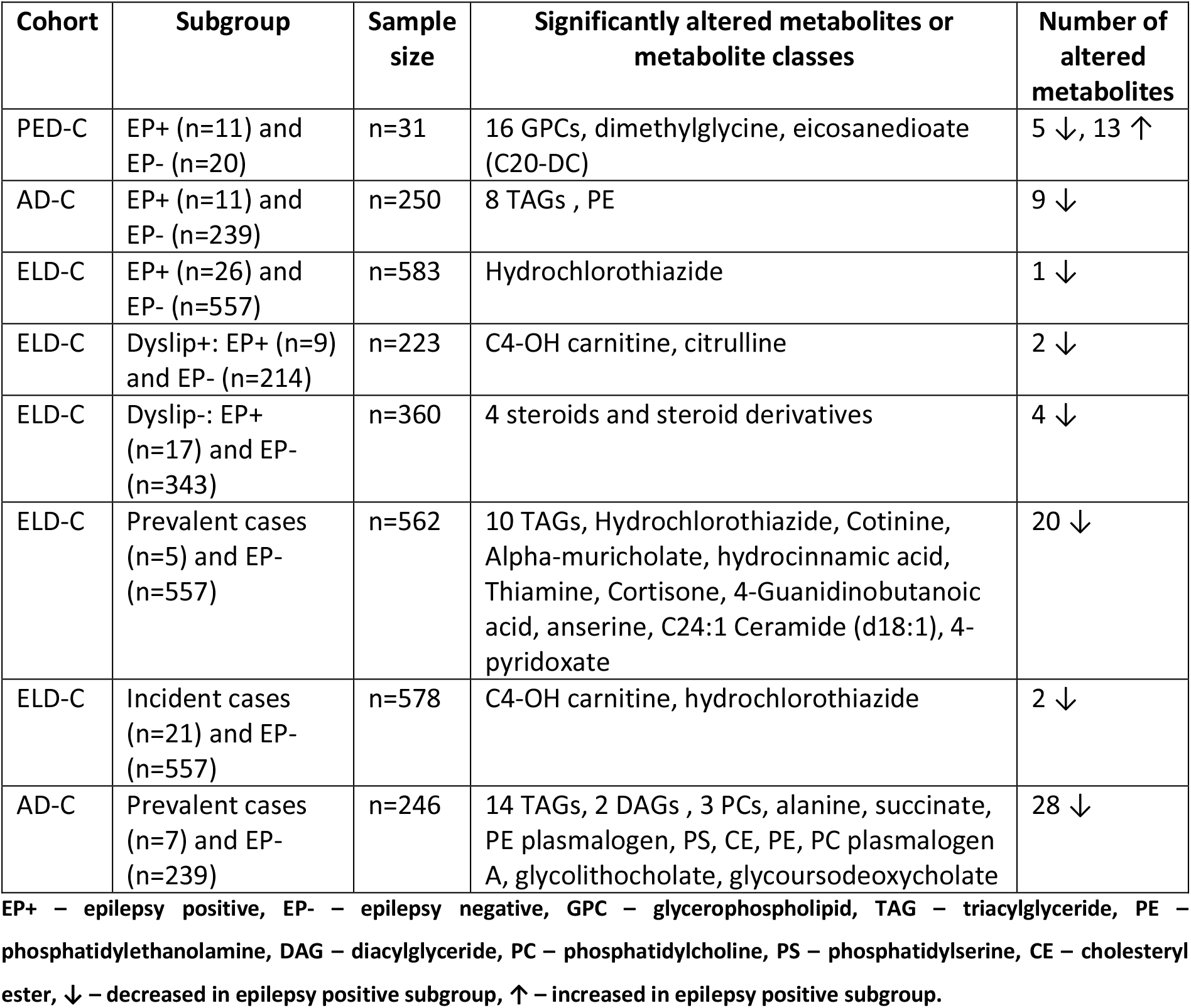
Summary of significantly altered metabolites in all analyzed cohorts and subgroups.

Pathway analysis revealed that among the metabolites that differ significantly between epilepsy-positive and epilepsy-negative patients in the PED-C, the lipid superpathway (p = 3.2*10^−4^), and phosphatidylcholine (p = 9.3*10^−8^) and lysophospholipid (p = 5.9*10^−3^) subpathways were statistically overrepresented. Analogously, in the AD-C, the triacylglyceride subclass was statistically overrepresented (p = 8.5*10^−5^) with the lipid superpathway (p = 1.4*10^−2^). The presented p-values are FDR-corrected.

The relatively large sample size of the ELD-C allowed for additional statistical tests in smaller subgroups (**Fig 1**). Firstly, we divided the cohort by the presence of dyslipidemia (E78) diagnosis and compared epilepsy-positive and negative cases within those subgroups. Among participants with dyslipidemia diagnosis, epilepsy was present in nine subjects out of 223, and we detected statistically significant differences in two metabolites (C4-OH carnitine and citrulline). In the subgroup without dyslipidemia, we detected statistically significant differences between epilepsy positive (17) and negative (343) cases in four metabolites (Chenodeoxycholate, Deoxycholate, Cortisone, and chenodeoxycholate/deoxycholate).

**Figure 1.**
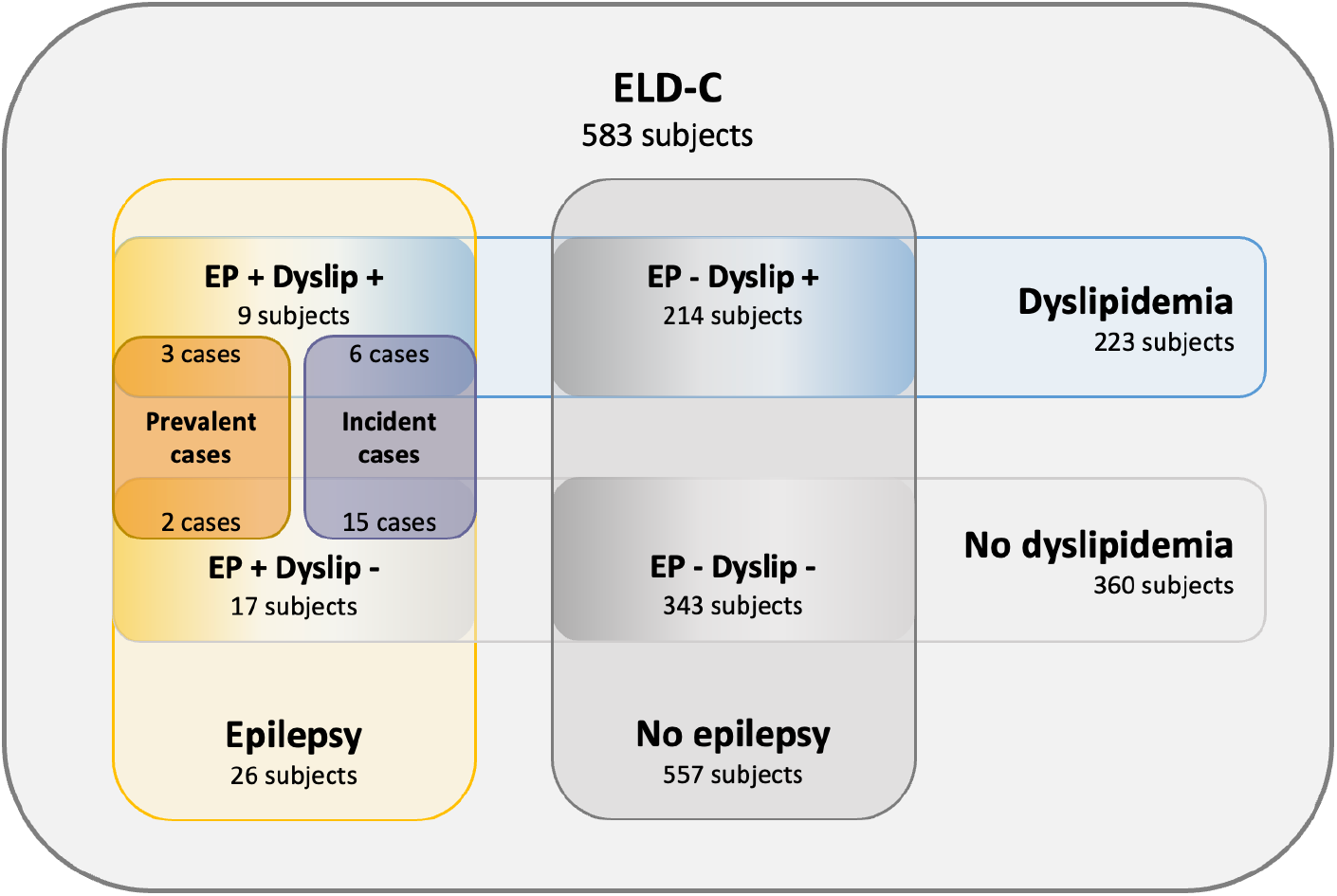
Subgroups in ELD-C. EP – epilepsy; Dyslip – dyslipidemia

Secondly, we divided the epilepsy-positive subgroup of the ELD-C into two subsets based on epilepsy onset before (prevalent cases) or after (incident cases) metabolomics sampling. Comparing incident cases with epilepsy negative subgroup revealed two statistically significantly altered metabolites – C4-OH carnitine and hydrochlorothiazide. However, when we compared the metabolic profile of prevalent cases with epilepsy negative subgroup, we saw significant changes in twenty metabolites, including multiple TAGs (**Table S2**).

We also reanalyzed the AD-C without the data from four epilepsy-positive patients with disease onset after metabolomics sampling. This analysis revealed 18 significantly altered metabolites belonging mainly to TAG, DAG, and GPL groups (**Table S2**).

## Discussion

We compared the metabolic profiles of individuals with and without epilepsy in three cohorts. Despite different patient ages, ages of disease onset, treatments with AEDs, and comorbidities, we observed changes in lipid metabolism in both pediatric and adult cohorts.

Lipids are heterogeneous organic compounds that fulfill various biological functions. They are essential structural components of biomembranes and serve as sources of heat and energy. They act as bioactive molecules and are essential in the metabolism of fat-soluble vitamins [^24,25^]. There are different ways of classifying lipids, but generally, they can be divided into five significant subcategories – fatty acids, triglycerides, phospholipids, sterol lipids, and sphingolipids [^25^]. Phospholipids include glycerophospholipids (GPLs) and phosphosphingolipids, which have great functional importance in the brain [^25^]. For example, GPLs are components of neuronal membranes and contribute to the formation of correct dendritic morphology [^26,27^]. The brain is the second most lipid-rich human organ; lipids comprise approximately half of its dry weight [^26^].

TAGs have a relatively minor role in brain lipid metabolism, but they act as a storage form of other lipid precursors and share a part of the GPLs synthesis pathway [^25^]. They are primarily synthesized in adipose tissue and the liver via the glycerol-3-phosphate pathway, where phosphatidate is one of the key intermediates. However, TAG synthesis also occurs in skeletal muscle, kidney, and brain [^25^].

Phospholipids are synthesized in the endoplasmatic reticulum of almost all cells except for erythrocytes, and this process is mainly active in the brain [^25^]. The main *de novo* synthesis pathways of all phospholipid classes and TAGs share common precursors, such as phosphatidate and diacylglycerol (**Fig 2**). Phosphatidate is a crucial intermediate in both pathways. Each phospholipid class has several synthesis pathways. For example, the *de novo* synthesis of phosphatidylcholines (PC) occurs via the Kennedy pathway, but PCs can also be synthesized by successive methylation of phoshpatidylethanolamines [^25^].

**Figure 2.**
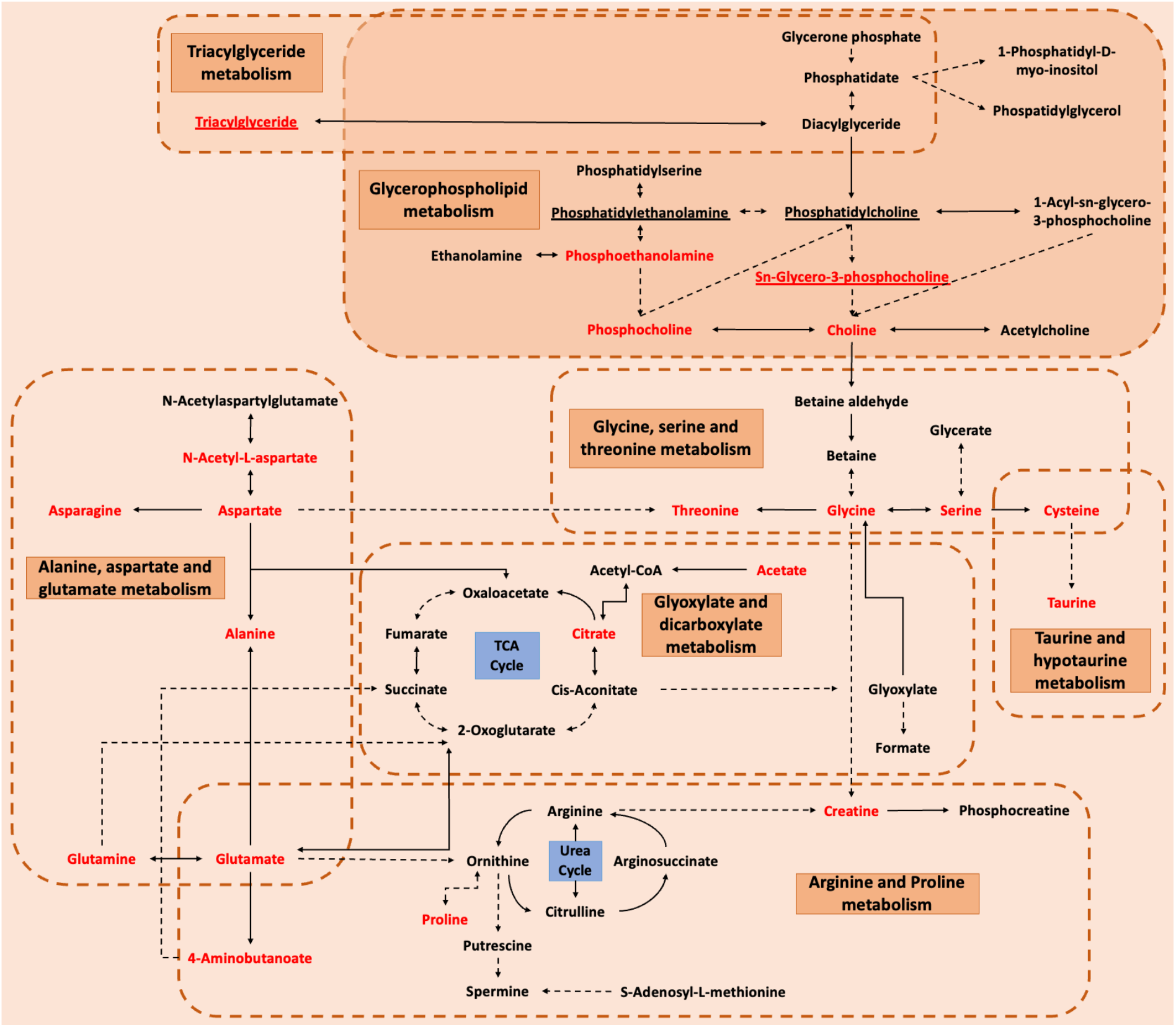
Main pathways and relevant metabolites involved in epilepsy. The dotted arrows mean multiple steps, while the solid arrows represent one step between two metabolites. The double-sided arrows indicate bidirectional, while the one-way arrows indicate unidirectional transformation. The red color emphasized the metabolites perturbed in both human and animal models. Orange dotted boxes and orange rectangles represent the main impacted metabolism pathways. Blue rectangles elucidate two crucial cycles, TCA (tricarboxylic acid) Cycle, and the Urea Cycle, linking up some metabolites. Alterations of underlined metabolites were observed in our cohorts. Figure adapted from Lai et al., 2022 [^13^].

Changes in lipid distribution (especially GPL and glycerolipid distribution) between the hippocampus and cortex, which potentially lead to loss of membrane integrity and perturbations in synaptic signaling, have been described in mouse models of epilepsy [^28^].

In our data, we observed changes in GPL metabolism in both pediatric and adult cohorts, although the exact metabolites did not overlap. Analyzing the elderly cohort as a whole did not reveal any significant changes in lipid metabolism. However, this could be due to only ∼20% of the epilepsy subgroup receiving their diagnosis before metabolomics sampling. Among participants who received their epilepsy diagnosis after sampling, the mean time from sample to diagnosis was nine years (SD=3.6) in ELD-C and 5.8 years (SD=2.9) in AD-C. The changes in metabolism may occur closer to the time of disease onset or after it. This hypothesis is supported by the changes in TAGs that we observed when we analyzed the subgroup of prevalent cases compared to the epilepsy-negative subgroup in ELD-C. Although there was no exact overlap, TAGs were also altered in AD-C, where the percentage of individuals with disease onset before sampling was much higher (∼64%). Alterations in TAGs have been described in animal models and patients with epilepsy [^13,29,30^]. However, the subgroups of epilepsy-positive subjects with disease onset before sampling were relatively small in both EstBB cohorts (AD-C and ELD-C). Therefore, further studies in cohorts with more epilepsy-positive cases must confirm this hypothesis.

Comparing the epilepsy incident cases with the epilepsy-negative subgroup in the ELD-C revealed lower C4-OH carnitine and hydrochlorothiazide. C4-OH carnitine is a short-chain acylcarnitine involved in beta-oxidation [^31^]. It can be a metabolite of lipid, amino acid, or carbohydrate metabolism [^32^]. It was altered in both the incident cases and the subgroup with dyslipidemia and epilepsy. However, the subgroups were relatively small, with six subjects belonging to both and accounting for two-thirds of the dyslipidemia-positive subgroup and approximately one-third of the incident cases. A higher level of C4-OH carnitine and other short-chain acylcarnitines would be expected in individuals with higher body fat and/or prediabetes [^33,34^]. Hydrochlorothiazide was also lower in incident cases compared to the epilepsy-negative subgroup of the ELD-C. However, we believe this is an incidental finding as hydrochlorothiazide is a diuretic commonly used to treat several diseases, including hypertension [^35,36^].

In the PED-C, most of the perturbed metabolites belonged to sn-glycero-3-phosphocholines. Alterations in this group have been described in previous studies in humans and animal models [^13,28,29,37–40^].

Most of the significant metabolites found in observed cohorts relate to the fluidity of cell membranes. It is known that the chemical structure of glycerophospholipids (GPLs) has a substantial role in the structure of the cell membrane. When the fatty acid chains of GPLs are saturated, they pack closer to each other, forming a more rigid structure and lowering the membrane’s fluidity. On the contrary, when GPL fatty chains are mono- or polyunsaturated, their non-polar intermolecular interactions weaken, packing becomes less dense, and the membrane becomes more fluid. In addition to GPLs, sterols (such as cholesterol) act as additional temperature-dependent membrane fluidity modulators. [^41^]

There are only a few previously published similar studies. The review by Lai et al. included 15 metabolomics studies of patients with epilepsy, nine of which used either serum or plasma samples. The number of patients with epilepsy included in the studies varied between 3 and 117. Some studies used healthy controls; others compared the metabolic profiles within epilepsy subgroups based on treatment, response to treatment, liver function, or other characteristics. [^13^] Most of these studies focused on small molecules and metabolites, such as glutamate, lactate, and citrate, which were disturbed in both patients with epilepsy and epileptic models. Only in one study were plasma lipidomics performed; however, the study group was very small (three individuals with epilepsy) [^42^].

This study was an exploratory analysis designed to generate hypotheses. Thus, limitations included the following. Firstly, the analyzed cohorts were very different, and there is a possibility of various confounding factors in each cohort. Secondly, although medications may affect the metabolomic profile, we needed more data to analyze their effect. The epilepsy-positive subgroups were small, and separating them into treated and untreated subgroups would render the sample size insufficient. In addition, the people who received treatment at the time of sampling used various medications. In the PED-C, we had information about all medications prescribed to them. However, for the AD-C and ELD-C, we only had data about medications belonging to ATC N03A, as we did not include any other medication groups in our data request. However, we do not have any reason to believe that the use of other medications would differ significantly between epilepsy-positive and negative subgroups within one cohort. Also, as dyslipidemia is a common disorder among the adult population [^43^], we were unable to rule out the presence of undiagnosed cases among the EstBB participants. Conversely, there might be participants with dyslipidemia who use statins or other lipid-modifying agents and therefore have normal blood lipid levels. Finally, we must consider that there might be misdiagnosis of epilepsy as well [^23^].

This study had several strengths as well. Using already existing data from the Estonian Biobank allows for identifying incident cases of epilepsy who are not receiving treatment yet. Also, to our knowledge, there have not been other studies of this scale that include lipidomics in the untargeted metabolomics analysis of individuals with epilepsy.

## Conclusions

Our results suggest that cell membrane fluidity may have a significant role in the mechanism of epilepsy, and changes in lipid balance may indicate epilepsy. However, further studies are needed to evaluate whether untargeted metabolomics analysis could prove helpful in diagnosing epilepsy earlier in cases where electroencephalogram is inconclusive.

## Acknowledgments

This work is supported by Estonian Research Council grants PRG471 and PRG1291. The Broad Institute Center for Mendelian Genomics (UM1HG008900) is funded by the National Human Genome Research Institute with supplemental funding provided by the National Heart, Lung, and Blood Institute under the Trans-Omics for Precision Medicine (TOPMed) program and the National Eye Institute. MHW is supported by K23HD102589. SP is supported by Estonian Research Council grant PSG774. DF and HP are supported by Estonian Research Council grants (PRG1095, PSG59, ERA-NET TRANSCAN-2 (BioEndoCar), Estonian Life Science Infrastructure for Biological Information (TT11)); Project No 2014-2020.4.01.16-0271, ELIXIR and the European Regional Development Fund through EXCITE Center of Excellence. We acknowledge the Estonian Biobank research team: Andres Metspalu, Lili Milani, Reedik Mägi, Mari Nelis, and Georgi Hudjashov, giving them credit for data collection, genotyping, QC, and imputation.

