## Supplemental Table 1 for "Untargeted metabolomics profiling in pediatric patients and adult populations indicates a connection between lipid imbalance and epilepsy"

**Table S1.** The clinical features, results of imaging and metabolic studies of epilepsy patients in the PED-C.

| PT | Sex | Age range (years) | OA (years) | Clinical characteristics, comorbidities | Epilepsy | EEG | Brain MRI | Treatment | Uniquely altered metabolites | Z-score |
|---|---|---|---|---|---|---|---|---|---|---|
| 1 | M | 11-15 | 0-5 | ID (severe), central hypotonia, dystonic movements | G40.2<br>G41.9 | Frontocentral epileptiform discharges in sleep | Normal brain structures, minimal enlargement of lateral ventricules | No treatment (Oxcarbazepine was started later) | 1-stearoyl-2-arachidonoyl-GPI (18:0/20:4) | -2.56 (< 2.5 pc) |
| 2 | M | 16-20 | 0-5 | ID (severe), brain structural anomaly, spasticity, weight and/or growth < -2.5 SD, microcephaly | G40.2 | Epileptiform discharges from occipital areas in sleep | Cavum septi pellucidi, cavum Vergae, pachygyria, hyperintense white matter signal in T1 and T2, calcifications | Lamotrigine<br>Valproic acid<br>Baclofen<br>Tizanidine | Valine<br>Phenylalanine | -2.32 (< 2.5 pc)<br>-2.91 (< 2.5 pc) |
| 3 | F | 11-15 | 0-5 | ID (profound), dysmorphic features, brain structural anomaly, central hypotonia, weight and/or growth<-2.5SD | G40.2<br>G40.4 | Multifocal epileptiform activity | Regional polymicrogyria, hypoplastic corpus callosum, cerebellar hypoplasia, large arachnoid cyst, subependymal heterotopias, calcifications | Vigabatrin<br>Lamotrigine | 2-aminoadipate<br>6-oxopiperidine-2-carboxylate | 3.26 (> 97.5 pc)<br>3.15 (> 97.5 pc) |
| 4 | M | 0-5 | 0-5 | ID (profound), dysmorphic features, brain structural anomaly, spasticity, microcephaly | G40.4 | Generalized and multifocal epileptiform activity | Biparietal schizencephaly, frontotemporal pachygyria/agyria, dysgenesis of corpus callosum, colpocephaly | Levetiracetam<br>Vigabatrin<br>Folic acid | No unique changes | |
| 5 | M | 6-10 | 6-10 | ID (severe), autism spectrum disorder, dysmorphic features, central hypotonia, coarctation of aorta. Clinically Kabuki syndrome | G40.3 | Epileptiform activity over the right hemisphere in sleep | Enlargement of the left lateral ventricle | Valproic acid | Asparagine<br>Indoleacetate<br>1-oleoyl-2-linoleoyl-GPE (18:1/18:2)*<br>1-stearoyl-2-arachidonoyl-GPE (18:0/20:4) | -2.37 (< 2.5 pc)<br>-2.46 (< 2.5 pc)<br>-2.71 (< 2.5 pc)<br>-2.12 (< 2.5 pc) |
| 6 | F | 16-20 | 11-15 | ID (profound), (auto)aggression, dysmorphic features, brain structural anomaly, weight and/or height < -2.5 SD, microcephaly | G40.2 | Epileptiform activity over the left hemisphere | Hypoplasia of corpus callosum, enlargement of lateral ventricules | Levetiracetam<br>Estradiol<br>Levothyroxin | N6-acetyllysine<br>5-hydroxylysine<br>Cystine<br>N-delta-acetylornithine<br>Myristoleoylcarnitine (C14:1)*<br>2-hydroxydecanoate<br>1-pentadecanoyl-2-docosahexaenoyl-GPC (15:0/22:6)*<br>Phosphatidylcholine (16:0/22:5n3, 18:1/20:4)* | 2.41 (> 97.5 pc)<br>2.00 (> 97.5 pc)<br>1.83 (> 97.5 pc)<br>2.05 (> 97.5 pc)<br>-2.53 (< 2.5 pc)<br>-1.81 (< 2.5 pc)<br>2.21 (> 97.5 pc)<br><br>2.41 (> 97.5 pc) |

| PT | Sex | Age range (years) | OA (years) | Clinical characteristics, comorbidities | Epilepsy | EEG | Brain MRI | Treatment | Uniquely altered metabolites | Z-score |
|---|---|---|---|---|---|---|---|---|---|---|
| | | | | | | | | | 1-margaroyl-2-arachidonoyl-GPC (17:0/20:4)*<br>Sphingomyelin (d18:1/18:1, d18:2/18:0)<br>Sphingomyelin (d18:1/20:1, d18:2/20:0)*<br>Sphingomyelin (d18:1/20:2, d18:2/20:1, d16:1/22:2)*<br>Sphingomyelin (d18:2/24:1, d18:1/24:2)*<br>Xanthine<br>Urate | 2.28 (> 97.5 pc)<br>2.08 (> 97.5 pc)<br>2.32 (> 97.5 pc)<br>1.96 (> 97.5 pc)<br>2.49 (> 97.5 pc)<br>2.52 (> 97.5 pc)<br>-15.95 (< 2.5 pc) |
| 7 | M | 11-15 | 0-5 | ID, autism spectrum disorder, brain structural anomaly | G40.3<br>G40.4 | Hypsarythmia | Cobblestone lissencephaly, subcortical white matter T2 hyperintense patches in frontal, parietal and occipital regions | Amantadine<br>Risperidon<br>Lamotrigine<br>Ethosuximide | Ethylmalonate<br>5alpha-androstan-3beta,17beta-diol disulfate<br>5alpha-androstan-3beta,17beta-diol monosulfate (2)<br>Androstenediol (3beta,17beta) disulfate (1)<br>Androstenediol (3beta,17beta) monosulfate (1)<br>2-aminooctanoate<br>3-carboxy-4-methyl-5-propyl-2-furanpropanoate (CMPF) | -1.91 (< 2.5 pc)<br>1.71 (>97.5 pc)<br>1.74 (>97.5 pc)<br>2.12 (>97.5 pc)<br>1.98 (>97.5 pc)<br>-2.34 (< 2.5 pc)<br>2.77 (>97.5 pc) |
| 8 | F | 11-15 | 0-5 | ID (severe), (auto)aggression, muscle weakness/hypotonia, ataxia, weight > 2.5 SD, height < - 2.5 SD, macrocephaly | G40.2<br>G40.8 | Multifocal epileptiform activity | Enlargement of lateral ventricules, cerebellar atrophy | Levetiracetam<br>Somatotropin | Bilirubin (E,E)*<br>16a-hydroxy DHEA 3-sulfate<br>1-stearoyl-2-dihomo-linolenoyl-GPI (18:0/20:3n3 or 6)* | 2.11 (>97.5 pc)<br>1.64 (>97.5 pc)<br>1.83 (>97.5 pc) |
| 9 | F | 0-5 | 0-5 | ID, (auto)aggression, autism spectrum disorder, dysmorphic features, brain structural anomaly, central hypotonia | G40.2 | Multiregional epileptiform activity dex>sin, epileptic encephalopathy | Temporomesial hippocampal sclerosis, slight hypomyelination, abnormal cerebral white matter | Diazepam<br>Sulthiame<br>Phenytoin | Homocitrulline<br>Myristate (14:0)<br>Palmitate (16:0)<br>Margarate (17:0)<br>Stearate (18:0)<br>Nonadecanoate (19:0)<br>Myristoleate (14:1n5)<br>10-heptadecenoate (17:1n7)<br>10-nonadecenoate (19:1n9)<br>Docosapentaenoate (n3 DPA; 22:5n3)<br>Hexadecadienoate (16:2n6)<br>(14 or 15)-methylpalmitate (a17:0 or i17:0)<br>(16 or 17)-methylstearate (a19:0 or i19:0)<br>Tetradecanedioate (C14-DC)<br>Hexadecanedioate (C16-DC)<br>1-stearoyl-2-docosapentaenoyl-GPC (18:0/22:5n3)*<br>1-oleoyl-2-docosahexaenoyl-GPC (18:1/22:6)*<br>1-linoleoyl-2-linolenoyl-GPC (18:2/18:3)*<br>1-stearoyl-2-oleoyl-GPI (18:0/18:1)<br>N-stearoyl-sphinganine (d18:0/18:0)<br>N-stearoyl-sphingosine (d18:1/18:0)<br>Myristoyl dihydrosphingomyelin (d18:0/14:0)<br>Sphingomyelin (d17:1/16:0, d18:1/15:0, d16:1/17:0)*<br>Sphingomyelin (d18:1/25:0, d19:0/24:1, d20:1/23:0, d19:1/24:0)*<br>Cholesterol<br>Carotene diol (1) | 5.74 (>97.5 pc)<br>2.29 (>97.5 pc)<br>1.93 (>97.5 pc)<br>2.38 (>97.5 pc)<br>2.41 (>97.5 pc)<br>3.04 (>97.5 pc)<br>2.47 (>97.5 pc)<br>2.66 (>97.5 pc)<br>2.08 (>97.5 pc)<br>2.41 (>97.5 pc)<br>2.17 (>97.5 pc)<br>2.74 (>97.5 pc)<br>2.63 (>97.5 pc)<br>3.47 (>97.5 pc)<br>2.65 (>97.5 pc)<br>1.85 (>97.5 pc)<br>1.96 (>97.5 pc)<br>1.98 (>97.5 pc)<br>1.99 (>97.5 pc)<br>2.06 (>97.5 pc)<br>2.11 (>97.5 pc)<br>1.73 (>97.5 pc)<br>1.96 (>97.5 pc)<br>1.82 (>97.5 pc)<br>3.35 (>97.5 pc)<br>2.18 (>97.5 pc) |

| PT | Sex | Age range (years) | OA (years) | Clinical characteristics, comorbidities | Epilepsy | EEG | Brain MRI | Treatment | Uniquely altered metabolites | Z-score |
|---|---|---|---|---|---|---|---|---|---|---|
| 10 | F | 11-15 | 0-5 | ID (severe), autism spectrum disorder, (auto)aggression, dysmorphic features, central hypotonia, weight and/or height < -2.5 SD, microcephaly | G40.2<br>G41.9 | No interictal epileptiform activity | Cavum septi pellucidi | No treatment | Lignoceroyl sphingomyelin (d18:1/24:0)<br>Sphingomyelin (d18:2/18:1)* | -2.36 (< 2.5 pc)<br>-1.96 (< 2.5 pc) |
| 11 | M | 6-10 | 0-5 | Speech developmental delay, brain structural anomaly. Seizure free for past 3 years. Variant in CSNK2A1 considered to be the cause of epilepsy | G40.3 | Frequent absences during wakefulness and light sleep, bilateral epileptiform activity interictally | Suspected malrotation of the left hippocampus | Lamotrigin<br>Topiramate<br>Clonazepam | 3-(3-hydroxyphenyl)propionate<br>3-hydroxyhippurate | 2.49 (>97.5 pc)<br>2.17 (>97.5 pc) |

PT – patient number, Age range – age range at sample collection, OA – age range at epilepsy onset, MRI – magnet resonance imaging, EEG – electroencephalogram, Treatment – medications used at sample collection, ID – intellectual disability, GPI – Phosphatidylinositol, GPE – Phosphatidylethanolamine, GPC – Phosphatidylcholine, * – the identification on metabolites marked with an asterisk is based on mass spectrometry data but no reference standards are currently available to verify the identity.

ICD-10 diagnosis codes: G40.2 – Localization-related (focal)(partial) symptomatic epilepsy and epileptic syndromes with complex partial seizures, G40.3 – Generalized idiopathic epilepsy and epileptic syndromes, G40.4 – Infantile spasms, G40.8 – Other epilepsy and recurrent seizures, G41.9 – Status epilepticus
