## Supplemental Table 2 for "Untargeted metabolomics profiling in pediatric patients and adult populations indicates a connection between lipid imbalance and epilepsy"

**Table S2.** Significant metabolic alterations in all cohorts and subgroups.

| Cohort | Subgroup | Sample size | Altered metabolite | Average Z-score | BH corrected p-value |
|---|---|---|---|---|---|
| PED-C | EP+ (n=11) vs EP- (n=20) | n=31 | 1-pentadecanoyl-2-arachidonoyl-GPC (15:0/20:4)* | 1.1310 | 0.0007 |
| | | | 1-stearoyl-2-docosapentaenoyl-GPC (18:0/22:5n6)* | -0.2727 | 0.0023 |
| | | | phosphatidylcholine (15:0/18:1, 17:0/16:1, 16:0/17:1)* | 1.4787 | 0.0024 |
| | | | 1-pentadecanoyl-GPC (15:0)* | 0.5531 | 0.0050 |
| | | | 1-myristoyl-GPC (14:0) | 0.4751 | 0.0050 |
| | | | Dimethylglycine | -0.7356 | 0.0050 |
| | | | 1-palmitoleoyl-GPC (16:1)* | 0.7933 | 0.0054 |
| | | | 1-pentadecanoyl-2-docosahexaenoyl-GPC (15:0/22:6)* | 1.6196 | 0.0059 |
| | | | 1-stearyl-2-linoleoyl-GPC (O-18:0/18:2)* | -0.4542 | 0.0068 |
| | | | 1-myristoyl-2-arachidonoyl-GPC (14:0/20:4)* | 1.0723 | 0.0068 |
| | | | 2-myristoyl-GPC (14:0)* | 0.3968 | 0.0069 |
| | | | 1-margaroyl-2-arachidonoyl-GPC (17:0/20:4)* | 0.8468 | 0.0070 |
| | | | 1-palmitoyl-2-palmitoleoyl-GPC (16:0/16:1)* | 1.7798 | 0.0123 |
| | | | 1-palmitoyl-2-eicosapentaenoyl-GPC (16:0/20:5)* | 1.9384 | 0.0181 |
| | | | 1-margaroyl-GPC (17:0) | 0.0360 | 0.0211 |
| | | | 1-myristoyl-2-palmitoyl-GPC (14:0/16:0) | 1.1674 | 0.0218 |
| | | | 1,2-dilinoleoyl-GPC (18:2/18:2) | -0.5755 | 0.0370 |
| | | | eicosanedioate (C20-DC) | -0.6091 | 0.0447 |
| AD-C | EP+ (n=11) vs EP- (n=239) | n=250 | C58:11 TAG | -0.3627 | 0.0036 |
| | | | C60:12 TAG | -0.3568 | 0.0056 |
| | | | C56:10 TAG | -0.3521 | 0.0058 |
| | | | C56:9 TAG | -0.4438 | 0.0118 |
| | | | C58:10 TAG | -0.4803 | 0.0184 |
| | | | C54:7 TAG | -0.4905 | 0.0199 |
| | | | C56:8 TAG | -0.5078 | 0.0232 |
| | | | C54:8 TAG | -0.4496 | 0.0215 |
| | | | C40:6 PE | -0.5396 | 0.0303 |
| ELD-C | EP+ (n=26) vs EP- (n=557) | n=583 | Hydrochlorothiazide | -0.2145 | 0.0046 |

| Cohort | Subgroup | Sample size | Altered metabolite | Average Z-score | BH corrected p-value |
|---|---|---|---|---|---|
| ELD-C | Dyslip+:<br>EP+ (n=9) vs<br>EP- (n=214) | n=223 | C4-OH carnitine<br>citrulline | -0.5301<br>-0.5002 | 0.0151<br>0.0094 |
| ELD-C | Dyslip-:<br>EP+ (n=17) vs<br>EP- (n=343) | n=360 | Chenodeoxycholate<br>Deoxycholate<br>Cortisone<br>chenodeoxycholate/deoxycholate | -0.4172<br>-0.4275<br>-0.2366<br>-0.4248 | 0.0007<br>0.0009<br>0.0121<br>0.0118 |
| ELD-C | Prevalent (n=5)<br>vs EP- (n=557) | n=562 | C43:2 TAG<br>Hydrochlorothiazide<br>Cotinine<br>Alpha-muricholate<br>C56:2 TAG<br>hydrocinnamic acid<br>Thiamine<br>C45:2 TAG<br>C54:1 TAG<br>C54:9 TAG<br>Cortisone<br>4-Guanidinobutanoic acid<br>C56:3 TAG<br>C43:1 TAG<br>C55:2 TAG<br>anserine<br>C24:1 Ceramide (d18:1)<br>C42:0 TAG<br>4-pyridoxate<br>C46:4 TAG | -0.5683<br>-0.2589<br>-0.2537<br>-0.3307<br>-0.5765<br>-0.6241<br>-0.1830<br>-0.4572<br>-0.5874<br>-0.3403<br>-0.2309<br>-0.1646<br>-0.6919<br>-0.4660<br>-0.6870<br>-0.3865<br>-0.7127<br>-0.3266<br>-0.2674<br>-0.5776 | <0.001<br><0.001<br><0.001<br><0.001<br>0.0081<br>0.0088<br>0.0114<br>0.0199<br>0.0359<br>0.0340<br>0.0410<br>0.0404<br>0.0381<br>0.0395<br>0.0379<br>0.0408<br>0.0472<br>0.0473<br>0.0452<br>0.0473 |
| ELD-C | Incident (n=21)<br>vs EP- (n=557) | n=578 | C4-OH carnitine<br>hydrochlorothiazide | -0.4748<br>-0.5372 | 0.0026<br>0.0023 |

| Cohort | Subgroup | Sample size | Altered metabolite | Average Z-score | BH corrected p-value |
|---|---|---|---|---|---|
| AD-C | Prevalent (n=7) vs EP- (n=239) | n=246 | C60:12 TAG | -0.4305 | <0.001 |
| | | | C58:11 TAG | -0.4370 | <0.001 |
| | | | glycoursodeoxycholate | -0.4493 | 0.0003 |
| | | | glycolithocholate | -0.5167 | 0.0036 |
| | | | C58:10 TAG | -0.5983 | 0.0048 |
| | | | C36:5 PC plasmalogen-A | -0.6655 | 0.0040 |
| | | | C56:9 TAG | -0.5212 | 0.0052 |
| | | | C54:7 TAG | -0.5849 | 0.0087 |
| | | | C56:8 TAG | -0.6096 | 0.0146 |
| | | | C40:6 PE | -0.6403 | 0.0135 |
| | | | C20:5 CE | -0.8176 | 0.0126 |
| | | | C56:10 TAG | -0.3889 | 0.0160 |
| | | | C34:0 PS | -0.8800 | 0.0160 |
| | | | C54:8 TAG | -0.5116 | 0.0181 |
| | | | C46:1 TAG | -0.4315 | 0.0280 |
| | | | C52:6 TAG | -0.4780 | 0.0324 |
| | | | C48:2 TAG | -0.4852 | 0.0406 |
| | | | C38:7 PE plasmalogen | -0.9199 | 0.0409 |
| | | | C34:2 DAG | -0.4578 | 0.0408 |
| | | | C52:7 TAG | -0.4636 | 0.0443 |
| | | | C38:6 PC | -0.8980 | 0.0431 |
| | | | succinate | -0.6033 | 0.0463 |
| | | | alanine | -0.6846 | 0.0488 |
| | | | C40:9 PC | -1.0107 | 0.0468 |
| | | | C58:9 TAG | -0.5655 | 0.0469 |
| | | | C40:6 PC | -0.7433 | 0.0512 |
| | | | C32:1 DAG | -0.4319 | 0.0498 |
| | | | C50:4 TAG | -0.4954 | 0.0487 |

EP – epilepsy, Dyslip – dyslipidemia, Prevalent – epilepsy diagnosed before sample date, Incident – epilepsy diagnosed after sample date, GPC – glycerophospholipid, DC – dicarboxylic acid, TAG – Triacylglyceride, PE – phosphatidylethanolamine, PS – phosphatidylserine, PC – phosphatidylcholine, CE – cholesteryl ester, * – the identification on metabolites marked with an asterisk is based on mass spectrometry data but no reference standards are currently available to verify the identity.
